# Using Project-based Learning to Enhance Curricular Integration and Relevance of Basic Medical Sciences in Pre-clerkship Years

**DOI:** 10.1101/2021.03.05.21252996

**Authors:** Fatma Alzahraa Abdelsalam Elkhamisy, Azza Hassan Zidan, Mohamed Fathelbab Fathelbab

## Abstract

**Introduction:** Integration levels vary in the basic medical pre-clerkship years. We aimed to discover the deficiencies and increase the level of integration for the multidisciplinary curriculum of the 1st 2 academic years at The Faculty of Medicine, Helwan University. We used Project-based learning (PtBL) via designing “Clinically-applied team-based integrated research project” tasks. The purpose was to make students fully appreciate the relationship between basic disciplines and their relevance to practice hence boosts their learning.

**Methods:** We designed interdisciplinary integrated research project tasks (cases/symptoms/signs) that followed each module’s objectives. Students worked in teams to write and deliver project reports. They analyzed the assigned tasks and used reasoning to create diagnoses, relate the condition to the disrupted normal structure/functions, suggest/contraindicate specific treatment plans, and create preventive plans based on their understanding of the basic medical sciences. A survey was introduced to assess students’ perceptions of the learning approach used. Students’ responses were analyzed.

**Results:** Deficient-, unrelated-, & should be related in an interdisciplinary way-topics in the curriculum were discovered and reported during the projects’ design. Students (n=694) completed the survey (52% response rate). Most (84.6%) were satisfied by the integrated interdisciplinary project, and 57.9% preferred substituting the traditional lectures completely by integrated PtBL. Students significantly (P=0.000) understood the relation between objectives of disciplines after project completion (mean 3.66, SD ±0.92) than before it (mean 3.46, SD ±0.91). A significant relation was detected between the rank given to the perceived degree of the integration between basic to clinical sciences in the projects and both the rank perceived for students’ developed clinical reasoning (P=0.000) and the students’ choice of implementing future learning via the integrated project tasks (P=0.002).

**Conclusions:** The PtBL can be used as a complementary learning method to elevate the level of integration within a multidisciplinary approach to boost students’ learning.

## Introduction

Medical schools are on different integration levels on Harden’s ladder.^1^ Many schools face a lower integration level in the basic medical preclinical years. Students also don’t fully appreciate the relevance of this academic phase to clinical practice due to little clinical data integration. The basic years’ curriculum in the Faculty of Medicine, Helwan University is mainly multidisciplinary with some interdisciplinary themes.

Project-based learning (PtBL) is inquiry-based learning in which learners assemble knowledge, comprehension, and prior experience. It starts with a problem & applied to small teams. Students’ activities are intended to construct a product not for “the studying itself”.^2^

To increase the level of integration in the basic years and increase the relevance of the curriculum for students; we adopted a project-based approach. We designed “Clinically-applied Team-based integrated research projects” tasks for students for 4 modules in the first 2 academic years. The level of integration in the tasks was interdisciplinary. Our purpose was to boost students’ learning in pre-clerkship years through enhancing the integration levels between basic sciences together, and between basic and clinical disciplines. Also, to evaluate and upgrade our curriculum to achieve a higher level of integration in the next years, based on the needs discovered while linking various disciplines’ objectives during the process of preparing these research project tasks. We assessed students’ perceptions of the approach used.

Medical education in Egypt is being reformed according to international standards. Our research results can be of help for any medical school that is still at a multidisciplinary level integration in the basic sciences in any national, regional, or international learning context.

## Materials and Methods

### The context

The study was carried out on phase 1 medical students (first- and second-preclinical years) in the Medical School of Helwan University, a public university in Egypt. It was carried out between February and June 2020. The first 2 years are mainly concerned with teaching basic medical sciences (pre-clinical, phase 1). We carried out our approach at the end of 2 modules in year 1; gastrointestinal (G.I.T.) and Locomotor, and 2 modules in year 2; Genitourinary and Blood/Endocrine. All lectures, practical lessons, problem-based learning (PBL) sessions were given online before assigning the projects’ tasks.

### Study design and tool used

This study employed a retrospective cross-sectional design to assess students’ perceptions of the approach used. A survey was prepared, sent to two experts in medical education for revision and validation, and then slight modifications were done based on their opinions. The survey was piloted on 30 students before dissemination. The survey consisted of a descriptive 16 item questionnaire regarding the students’ perceptions on the impact of the PtBL approach used. Answers were collected by using yes/no questions and a five-point Likert scale (1; extreme negative response, and 5; extreme positive response).^3^ Adding open-ended written feedback was optional for which breakdown analysis was done to analyze the written data. The survey items were internally consistent as the calculated Cronbach alpha for the close-ended questions was 0.77.

### Sample

A convenience sample was done as students filled in the survey according to their willingness to do so. The total number of students on which the survey was distributed was 1334. Inclusion criteria were being a current first- or second-year medical student in the faculty that completed the required tasks and agree to participate in the survey.

### Implementation

The authors volunteered to review the modular curricula and design **a wide variety of** interdisciplinary integrated project’ topics (cases/symptoms/signs) that followed each module’s objectives (40-65 tasks/module). Disciplines’ instructors modified the tasks to suit the intended learning outcomes.

Each research project topic was a designed case/symptom/sign that integrates all disciplines represented in the module. Students were requested to discuss with each other, search through their books, previously studied material, and online to write an integrated report on the task. Students had to analyze the topic of the task and use clinical reasoning to create a reasonable differential diagnosis and/or a provisional one based on their understanding of the sciences of diseases in the preclinical phase (e.g. pathology). The students are required to discuss and relate the condition to the normal bases that were disrupted (i.e. anatomy, histology, physiology, and biochemistry). Besides; they had to discuss/suggest/contraindicate specific treatment plans (i.e. pharmacology), as well as create a preventive plan (i.e. epidemiology) whenever possible.

Students were divided into teams each is formed of 5 members. The evaluation rubrics and the project requirements were clear to students and evaluators at the beginning of implementation. Instructors responded to the students’ inquires. Students uploaded their finished tasks to their learning management system (LMS) before the announced due dates.

An online survey prepared on Google Forms was introduced after the due date of the projects’ delivery to assess students’ perception of the new intervention. Students were asked to voluntarily participate in the online survey administered on their Learning management system.

### Data Collection

The survey data were collected on Excel spreadsheets. Data about missing topics that need to be added in the modular curriculum and topics that can be involved in more interdisciplinary themes were collected and written in a report during the process of the curricular review for tasks’ design.

### Statistical analysis

The survey data were analyzed using IBM-SPSS (Statistical Package for Social Sciences) Version 25.0; a Chi-square test was used to compare qualitative variables, and t-tests were used to compare quantitative variables. Z-tests were used to compare proportions. The level of significance was P <0.05. Cronbach alpha test was used to analyze the internal consistency of the survey items and a value > 0.7 is considered internally consistent.

### Ethical approval

The study complied with Helwan University Ethics Committee Guidelines for research with humans and has been approved by the Helwan University Ethics Committee (Serial number: 48-2020), organized, and operated according to the declaration of Helsinki 1975. Consent of participation was obtained from all participants before they participated in the study, participants acknowledge that they cannot be identified via the paper; and that they have been fully anonymized in the research.

## Results

Some deficient basic sciences topics in the modular curricula which need to be added by the disciplines to complete a picture relevant to the clinical application were discovered during the projects’ topics design as well as reported by students in their written feedback. Also, some objectives were being educated separately although they can be related to other disciplines in an interdisciplinary approach. A report was written for deficiencies and suggestions to be discussed, modified, and implemented in the future curriculum.

Regarding the survey responses, 694 students of phase1 (first and second year) medical students participated in the study. They represented 52% of the total phase 1 students. Students’ age range was between 17-19 years. Demographic data of study participants are shown in table (1).

The majority of students (84.6%, n=587) preferred doing one integrated interdisciplinary PtBL task instead of multiple separate tasks in all disciplines. Students appreciated the connection between disciplinary objectives of their tasks higher (mean 3.66, SD ± 0.92) after doing the PtBL required tasks compared to before it (mean 3.46, SD ± 0.91), (P=0.000). Level of integration between disciplines in the tasks varied in the 4 modules; the very good & excellent ranking for integration between basic sciences ranged between 35.2% (n=244) to 85.9% (n=596) and for basic/clinical integration between 33.3% (n=231) to 78.8% (n=547). Overall, most students (60.6%, n=420) ranked their appreciation of the relationship between all basic sciences in each research topic by ranks 4 and 5, and 56.1% (n=389) ranked the relation between basic and clinical sciences through the integrated researches by ranks 4 and 5 (Tables 2 & 3).

**Table 1:** Demographic data of study participants.

| **Character** | **No.** | **%** |
| --- | --- | --- |
| **Academic year** |  |  |
| First | 408 | 58.8 |
| Second | 286 | 41.2 |
| **Gender** |  |  |
| Male | 361 | 52 |
| Female | 333 | 48 |
| **High school background** |  |  |
| Egyptian high-school education (Thanaweya Amma) | 410 | 59.1 |
| IGCSE | 52 | 7.5 |
| American diploma | 17 | 2.4 |
| Stem | 13 | 1.9 |
| Arabic non-Egyptian | 193 | 27.8 |
| Other international certificates | 9 | 1.3 |
| **Academic performance (Grade)** |  |  |
| A | 269 | 38.8 |
| B | 182 | 26.2 |
| C | 86 | 12.4 |
| D | 34 | 4.9 |
| F | 56 | 8.1 |
| Postponed previous exams | 67 | 9.7 |
| **Family permanent residency** |  |  |
| In Egypt | 492 | 70.9 |
| Outside Egypt | 202 | 29.1 |
| **Total** | **694** | **100** |

No.: Number

**Table 2:** Students’ responses to the survey questions measuring their learning preferences regarding the Project-based learning approach used.

| **Survey item** | **No.** | **%** |
| --- | --- | --- |
| **Do you prefer doing separate researches in each subject or one integrated research as you did?** |  |  |
| Separate researches | 108 | 15.6 |
| One integrated research | 586 | 84.4 |
| **Do you prefer learning by explaining as in usual lectures or by explaining cases with all topics integrated like in the research?** |  |  |
| usual lectures | 293 | 42.2 |
| explaining in cases | 401 | 57.8 |
| **Did you prefer to do the research individually or in a team?** |  |  |
| Individually | 137 | 19.7 |
| In a team | 557 | 80.3 |
| **In your opinion, what was the suitable number of students that should form each team?** |  |  |
| 1 | 137 | 19.7 |
| 2 | 5 | 0.7 |
| 3 | 23 | 3.3 |
| 4 | 36 | 5.2 |
| 5 | 450 | 64.8 |
| 6-10 | 40 | 5.8 |
| other | 3 | 0.3 |
| **Generally speaking, do you suggest changing scoring of some tests/assignments as pass/fail instead of discriminative ranks?** |  |  |
| Yes | 392 | 56.5 |
| No | 302 | 43.5 |
| **In future years, do you recommend making a short presentation by each team for the integrated research to be discussed in front of all other students to expand the benefit?** |  |  |
| Yes | 414 | 59.7 |
| No | 280 | 40.3 |
| **In case integrated researches were used as a future method of learning, select most suitable approach** |  |  |
| For revision at the end of the module | 163 | 23.5 |
| Each case become the theme of the week and all subjects discuss it | 203 | 29.3 |
| As an assignment during the module | 122 | 17.6 |
| Cases discussed in integrated lectures | 148 | 21.3 |
| As part of the portfolio cases for self-learning | 44 | 6.3 |
| Other (please specify) | 14 | 2.0 |
| **Total** | 694 | 100.0 |
| No.: Number |  |  |

Most students (57.5%, n=399) ranked their developed reasoning skills by the PtBL experience as ranks 4 and 5. Results were significantly related to the perceived degree of basic and clinical sciences integration in the tasks (P=0.000), (Tables 3 & 4).

**Table 3:** Students’ ranked perceptions of the Project-based learning approach used.

| Survey Item | Rank* |  |  |  |  |  |  |  |  |  |
| --- | --- | --- | --- | --- | --- | --- | --- | --- | --- | --- |
|  | 1 |  | 2 |  | 3 |  | 4 |  | 5 |  |
|  | No. | % | No. | % | No. | % | No. | % | No. | % |
| Developed student`s clinical reasoning skills | 21 | 3.0 | 70 | 10.1 | 204 | 29.4 | 256 | 36.9 | 143 | 20.6 |
| The appreciated relation between basic sciences & their clinical application | 17 | 2.4 | 62 | 8.9 | 226 | 32.6 | 256 | 36.9 | 133 | 19.2 |
| The appreciated relation between all basic sciences | 15 | 2.2 | 47 | 6.8 | 212 | 30.5 | 286 | 41.2 | 134 | 19.3 |
| Developed research skills | 6 | 0.9 | 8 | 1.2 | 166 | 23.9 | 369 | 53.2 | 145 | 20.9 |
| Developed team work skills | 57 | 8.2 | 22 | 3.2 | 106 | 15.3 | 244 | 35.2 | 265 | 38.2 |
| Satisfaction on changing the scoring system to pass/fail instead of discriminative ranks | 31 | 4.5 | 42 | 6.1 | 161 | 23.2 | 187 | 26.9 | 273 | 39.3 |
| The degree of integration between the disciplines involved in the students` tasks before doing the task | 17 | 2.4 | 69 | 9.9 | 268 | 38.6 | 258 | 37.2 | 82 | 11.8 |
| The degree of integration between the disciplines involved in the students` tasks after finishing the task | 14 | 2.0 | 49 | 7.1 | 221 | 31.8 | 286 | 41.2 | 124 | 17.9 |

No.: Number; Rank: 1 (strongly disagree/least negative response), 5 (strongly agree/maximum positive response). 4 and 5 are considered positive results

**Table 4:** The relationship between students’ rank of appreciated link of basic to clinical and the perceived rank for developed clinical reasoning skill.

|  |  |  | Rank of developed clinical reasoning skill |  |  |  |  |  |
| --- | --- | --- | --- | --- | --- | --- | --- | --- |
|  |  |  | 1 | 2 | 3 | 4 | 5 | Total |
| Students' rank of appreciated link of basic to clinical | 1 | No. | 6 | 7 | 2 | 1 | 1 | 17 |
|  |  | % | 0.9% | 1.0% | 0.3% | 0.1% | 0.1% | 2.4% |
|  | 2 | No. | 7 | 25 | 15 | 14 | 1 | 62 |
|  |  | % | 1.0% | 3.6% | 2.2% | 2.0% | 0.1% | 8.9% |
|  | 3 | No. | 3 | 23 | 124 | 63 | 13 | 226 |
|  |  | % | 0.4% | 3.3% | 17.9% | 9.1% | 1.9% | 32.6% |
|  | 4 | No. | 3 | 14 | 56 | 140 | 43 | 256 |
|  |  | % | 0.4% | 2.0% | 8.1% | 20.2% | 6.2% | 36.9% |
|  | 5 | No. | 2 | 1 | 7 | 38 | 85 | 133 |
|  |  | % | 0.3% | 0.1% | 1.0% | 5.5% | 12.2% | 19.2% |
| Total |  | No. | 21 | 70 | 204 | 256 | 143 | 694 |
|  |  | % | 3.0% | 10.1% | 29.4% | 36.9% | 20.6% | 100.0% |
| P=0.000 |  |  |  |  |  |  |  |  |

No.: Number; Rank: 1 (strongly disagree/least negative response), 5 (strongly agree/maximum positive response). 4 and 5 are considered positive results

Most of the students (76.2%, n=529) perceived their level of developed teamwork skills by the PtBL tasks used as ranks 5 and 4. The majority of students (80.4%, n=558) preferred to do the task in a team. On a range of 1 to 10 members; most students preferred teams formed of 5 members (64.9%, n=452). Also, most students (74.1%, n=514) ranked their perceived level of developed research skills as ranks 4 and 5 (Tables 2 & 3).

Most students (59.5%, n=413) recommended making a short presentation by each team for the prepared PtBL task to be discussed in front of all other students to expand the benefit. They preferred learning by integrated cases with all topics included like the PtBL experience (57.9%, n=402) over the traditional disciplinary lectures (Table 2).

Students’ acceptance to future use of this PtBL was significantly related to their esteemed degree (i.e. ranking) of integration between basic and clinical sciences in the tasks (P=0.002) (Table 5), but not to the degree of integration between basic sciences together.

**Table 5:**
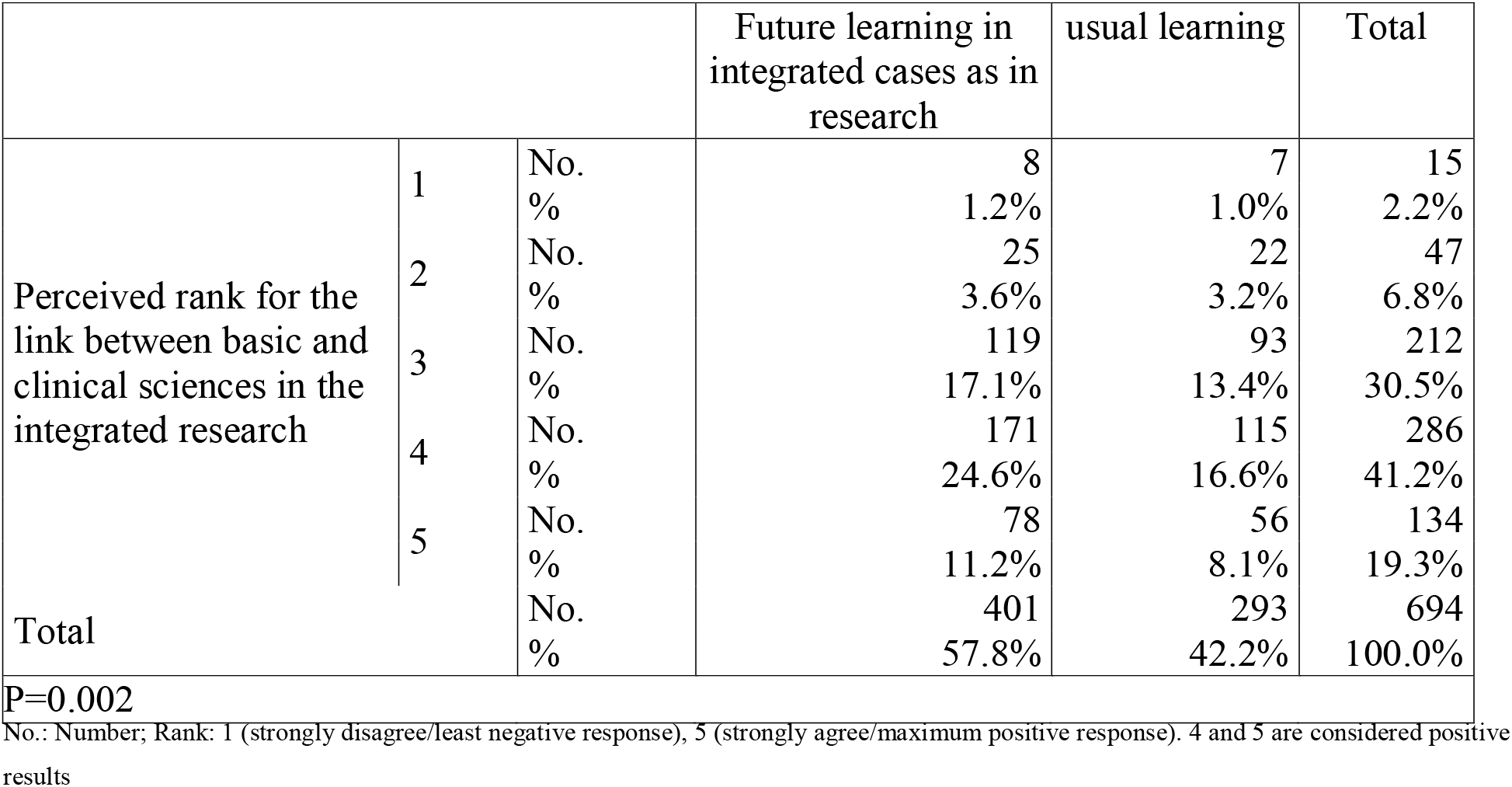
The relationship between students’ responses regarding future learning by the integrated researches method and the degree of perceived integration between basic and clinical sciences in the case.

Most students (66.2%, n=459) were satisfied by ranks 4 and 5 on changing the scoring system to pass/fail instead of discriminative ranks in evaluating the projects. They (56.6%, n=393) agreed to future changing the scoring of some tests/assignments as pass/fail instead of discriminative ranks (Tables 2 & 3).

Methods of future learning through the interdisciplinary PtBL selected were; making these projects themes for learning weeks; with all disciplines discuss a task each week (29.3%, n=203). Also, assigning these tasks at the end of the module as a method for concluding work and revision (23.5%, n=163). Some students (21.3%, n=148) preferred the discussion of the topics in integrated lectures (21.3%, n=148). Others (17.6%, n=122) preferred making the integrated interdisciplinary tasks assignments during the modules (Table 2).

## Discussion

In our study, we implemented PtBL by assigning “clinically-applied team-based integrated research project tasks” students in the pre-clerkship years studying basic medical sciences. A cohort of 694 students shared their perceptions of the PtBL used. Most were satisfied by implementing the one integrated interdisciplinary research project task, team-based work with 5 members in each team, and the pass/fail scoring system. A significant relation was detected in the students’ ranking of the relation between the task’s disciplinary objectives after completing it compared to before it, and between the integration with clinical sciences on one side, and both the developed clinical reasoning and the students’ choice of implementing future teaching via the integrated project tasks on the other side.

Project-based learning (PtBL) is an inquiry-based approach based on constructivism theory.^2^ It is sometimes confused with problem-based learning (PBL) and even some put the same abbreviation PBL for both of them. Both are applied to small students’ groups, and both start with a problem. However, the main focus of the PtBL approach is making learners construct a product while the problem-based learning focus is to make learners study. In PtBL, the teacher role is only advisory when needed not a facilitating role present in the whole session like in PBL. So, learners in PtBL have more control over the learning process. Problem-based learning also has a certain method/steps of application that require to be followed while PtBL is more flexible.^4^

Our approach proved helpful for learning. Project-based learning research experience is well perceived by students in other studies.^5^ In medical education, it was also used to increase medical students’ empathy, teach medical bioethics, and microbiology.^6–8^

Harden (2000)^1^ proposed 11 levels of integration simulating the steps of the ladder. The higher the ladder step, the higher the integration level of the curriculum. He describes the multidisciplinary approach of integration (Step 9) as bringing disciplines together around themes. He proposed that themes can be certain clinical conditions or body systems. Interdisciplinary integration (Step 10) is a higher level of integration in which no discipline boundaries are obvious. Disciplines melt in each other in a course. Multidisciplinary curricula that are organized around body systems require effort to organize the disciplines within each system in a careful way to avoid reverting to a lower level of integration inside the system. For example, in the G.I.T. body system, disciplines should be organized around the esophagus, around the stomach, around the liver, etc. Teaching every discipline on its own within the body system without good organization with other disciplines decreases the level of integration. Multidisciplinary curricula differ in the organization of their disciplinary content. Melting disciplines together in projects inside a multidisciplinary curriculum takes a further step toward interdisciplinary integration.

Based on this experience, the degree of integration within the body system modules was variable from one module to another. This affected the degree of integration within our project tasks. Students’ ranking of the level of integration achieved between all basic sciences together, and between basic and clinical sciences in the objectives of each research topic was 4 and 5 in 60.6%, and 56.1% respectively as perceived by students. These objectives were following the preset curricular objectives. Although our curriculum follows a multidisciplinary approach; the relation between disciplines was not that obvious for students. Curriculum revision is needed to adjust the preset objectives and increase the integration level both vertically and horizontally.

Potentiating the link between basic sciences together on one hand, and between the basic and clinical sciences, on the other hand, helps develop clinical reasoning skills in students starting from basic sciences years.^9^ Medical students are aware of the aim of the educational reform and are more willing to higher levels of integration; they preferred doing one integrated research task in each module instead of multiple separate tasks in each discipline. The perceived degree of integration between disciplines also significantly affected the perceived levels of developed clinical thinking (reasoning) skills. Other authors concluded similar results.^10^

Percentage of students preferring future learning through cases with all topics integrated like in the researches instead of the multidisciplinary learning was significantly related to increasing the level of integration between basic and clinical sciences. Similarly, Senti et al.^11^ showed the increased students’ interest, acquired knowledge, and skills of adopting basic/clinical integrated programs.

Semin et al. (2018) implemented multidisciplinary case-based small group discussions to integrate basic medical sciences with clinical situations for 39 students in 5 body systems and found that 62% of students find the integrated cases useful for their learning.^12^ They carried out discussions on 3 hours sessions and without enough prior knowledge for students and set the outcomes for each case. Our study was carried out on a larger number of participants (694), and students were the main drivers of their learning process, and they had previous knowledge from all disciplines and we derived the outcomes from the curriculum that is required to be evaluated and improved to report the deficiencies present and avoid confusing students with them.

Yune & Jung (2018)^13^ showed that students’ academic performance significantly increased on doing a curricular revision that enhanced integration between basic & clinical sciences in the pre-clerkship years. Deeper understanding with easier retrieval and transfer of basic medical knowledge happens better when linked to the clinical context.^14^ This is also supported by the adult learning theory which points that adults are most engaged in learning subjects with immediate practical relevance.^15^

Majority of students preferred to do the tasks in a team rather than individually. Five-members were the most reported suitable number for building a team. This number proved helpful in developing the academic research skills for postgraduates also.^16^ The nature of scientific material educated seems to potentiate their tendency for teamwork.^17^ Explaining the basics of effective teamwork at the students’ admission and augmenting the clinical application in curricula may be considered.

Most students showed positive attitudes towards changing the scoring system to pass/fail instead of discriminative ranks. Moreover, most students agreed to the suggestion of future changing the scoring of some tests as pass/fail instead of discriminative ranks. The pass/fail scoring system resulted in a good performance and great satisfaction in other preclinical medical courses.^18^ It exerts positive influences on learning by supporting students’ psychological health and wellbeing.^19^

Our study limitations include that no control group with academic performance was done, being carried out in a single institution.

## Conclusion

The interdisciplinary PtBL can be used to enhance the integration level between disciplines in pre-clerkship medical years. Advantages include emphasizing the integration between basic sciences together and with clinical application, developing research, team-work, and clinical reasoning skills which are needed for future medical practice. The team-based integrated research project tasks and the pass/fail scoring system are well perceived by students. Students’ acceptance to future use of the interdisciplinary PtBL is related to their esteemed degree of integration between basic and clinical sciences in the projects’ tasks. The process of preparation of the tasks helps the staff evaluate the curriculum and discover areas that need modifications for a higher level of integration.

## Data Availability

The datasets generated and analyzed during the current study are available at the corresponding author upon request.

## Acknowledgements

The authors would like to thank all the management & staff members, and the students at Helwan Medical School who supported and/or participated in implementing the experience and/or shared their perceptions.

